# HIV Remains a Risk Factor for Unfavorable Tuberculosis Treatment Outcomes in the Era of Universal Access to Antiretroviral Therapy in Botswana

**DOI:** 10.1101/2025.11.26.25340699

**Authors:** Azadeh Baniasad, Siheng Chao, James A. Nguyen, Emily Tian, Chawanga Modongo, Volodymyr M. Minin, Jessalyn N. Sebastian, Sanghyuk S. Shin

**Affiliations:** Department of Statistics and Data Science, University of California, Los Angeles; Department of Statistics, University of Pittsburgh; Department of Mathematics, University of California, Irvine; Department of Statistics and Applied Probability, University of California, Santa Barbara; Victus Global Botswana Organisation, Gaborone, Botswana; Department of Statistics, University of California, Irvine; Sue & Bill Gross School of Nursing, University of California, Irvine; Division of Pulmonary, Allergy, Critical Care, and Sleep Medicine, Department of Medicine, University of Pittsburgh

## Abstract

Botswana implemented its universal “Treat All” antiretroviral therapy (ART) policy in 2016, expanding treatment eligibility to all people living with HIV (PLHIV). HIV has been known to be a leading risk factor for tuberculosis (TB) and poor TB treatment outcomes. The primary goal of this study is to assess whether HIV infection and HIV-associated immunosuppression remain risk factors for unfavorable TB treatment outcomes in the Post–Treat All era. We analyzed 636 TB patients treated in Gaborone (2017–2023), of whom 54.4% were HIV-positive. Unfavorable outcomes (death, failure, or loss to follow-up) occurred in 19.7% of HIV-positive and 8.5% of HIV-negative patients. We used logistic regression to estimate unadjusted and covariate-adjusted associations between TB treatment outcome and HIV status and between TB treatment outcome and CD4+ T-cell count. HIV-positive patients had 2.5-fold higher odds of unfavorable outcomes compared with HIV-negative patients [adjusted OR: 2.51, 95% CI: (1.48, 4.38)], controlling for age, sex, TB history, distance to clinic, substance use, and occupational status. PLHIV with CD4+ T-cell < 200 cells/*µ*L was associated with approximately three-fold higher odds of unfavorable outcomes compared with HIV-negative participants [OR: 3.12, 95% CI: (1.65, 5.97)]. The secondary goal was to test whether the HIV effect changed following Treat All implementation. We combined the data from 2017–2023 with a Pre–Treat All cohort (2012–2016, n= 233, HIV prevalence 60.8%) and fit a frequentist logistic regression and Bayesian mixed-effects models with an interaction term that allows treatment era (Pre- vs. Post-Treat All) to modify the effect of HIV on TB treatment outcome. The estimated change in the HIV effect was uncertain [relative OR: 0.41; 95% CI: (0.11, 1.55)]. Combining the two Botswana data sets with 12 Pre- and Post-Treat All studies from neighboring Ethiopia showed that the pooled effect of HIV infection on unfavorable TB outcome has increased in the Post-Treat All period [relative OR: 2.39; 95% BCI: (1.36, 3.34)].

## 1 Introduction

Tuberculosis (TB), a disease caused by the airborne pathogen *Mycobacterium tuberculosis* (Mtb), is one of the top 10 leading causes of death in the world according to the World Health Organization (WHO). According to the World Health Organization (WHO), tuberculosis (TB) caused an estimated 1.25 million deaths in 2023 (World Health Organization, 2024b). Although TB mortality rate has steadily declined since the 1950s, it has recently seen significant resurgence since 2020 (for Disease Control and Prevention, 2024). The most widely used TB treatment involves over six months of intensive combination chemotherapy, which is able to fully cure the patient.

Human immunodeficiency virus (HIV) infection is the most well-known risk factor for developing active TB. People living with HIV (PLHIV) are 15-21 times more likely to develop active TB compared to those who test negative for HIV (Buziashvili et al., 2024; World Health Organization, 2024a). In 2023, PLHIV accounted for approximately 161,000 of the 1.25 million TB deaths (World Health Organization, 2025b). HIV weakens the immune system by targeting CD4+ T-cells, which are essential components of the adaptive immune system in combating pathogens such as Mtb. Consequently, PLHIV have historically been more vulnerable to Mtb infections and have been at a higher risk for unfavorable TB treatment outcomes. HIV status and severity can be assessed by circulating CD4+ T-cell counts measured via flow cytometry. Antiretroviral therapy (ART) is a treatment for HIV, which includes a family of medications designed to inhibit the HIV viral cycle within lymphocytes. This diminishes the presence of HIV in the body and keeps the immune system functional by retaining healthy CD4+ T-cell counts. Clinical trials have shown association between early ART therapy and reduced risk from adverse events from TB (Havlir et al., 2011).

In 2016, the government of Botswana announced its universal ART policy, which guarantees access to ART for all PLHIV, regardless of CD4+ T-cell counts. Since then, Botswana has surpassed the UNAIDS gold standard of 95-95-95: 95 percent of PLHIV are aware of their status, 95 percent of those diagnosed PLHIV receive ART, and 95 percent of those on ART achieve viral suppression. Recent studies have already found an association between universal ART coverage and viral suppression (Makhema et al., 2019). Before implementation of Botswana’s 2016 universal ART policy (Pre-Treat All), a study conducted in Gaborone as part of the Kopanyo project examined demographic and clinical factors associated with TB treatment outcomes among people with and without HIV (Zetola et al., 2016). The time period for this Pre-Treat All study stretched from 2012 to early 2016. Using these data, Shin et al. (2018) identified a positive association between the presence of HIV infection and the unfavorable TB treatment outcomes. Our primary aim was to assess whether HIV infection remains a risk factor for unfavorable TB treatment outcomes in the Post–Treat All era, and our secondary aim was to determine whether this association changed compared to the Pre–Treat All period.

## 2 Methods

### 2.1 Setting

Botswana has one of the highest TB incidences worldwide, with an estimated prevalence of around 244 cases per 100,000 people in 2023. It also has a significant population of PLHIV, at around 17 percent of the adult population. Among TB patients in Botswana, the HIV co-infection rate is around 44 percent (Centers for Disease Control and Prevention, 2025; Zetola et al., 2021). The metropolitan area of Gaborone accounts for more than 10 percent of Botswana’s population and serves as the political and cultural center of the country.

### 2.2 Data Source

The datasets for the Pre-Treat All and Post-Treat All studies were derived by combining surveys, clinical biomarkers, and data from the electronic tuberculosis registry of the Botswana Ministry of Health. Additional details about patient enrollment and sample collection can be found in a previously published article (Shin et al., 2018). According to WHO guidelines, a favorable treatment outcome describes a TB patient who has completed treatment or experienced a full recovery, while an unfavorable treatment outcome refers to a TB patient who did not complete treatment, died, or is lost to follow-up. The WHO categorizes patients into normal (count *≥* 500 cells/*µ*l), compromised (200*≤* count < 500 cells/*µ*l), or severely compromised (count < 200 cells/*µ*l) levels of CD4+ T-cell counts; we used these guidelines to partition our HIV-positive patients into one of the three groups (World Health Organization, 2025a). Patients who drink and/or smoke were classified as individuals with substance use. Both students and working individuals were grouped together as individuals having occupation.

We also performed a Bayesian meta-analysis that incorporated data from 12 other studies conducted in Ethiopia. Ethiopia, like Botswana, is a Sub-Saharan African country with a high burden of TB and HIV (Bagcchi, 2023). Ethiopia also implemented a similar “test and treat” policy for HIV in 2016 (Bantie et al., 2022). These studies were found through a literature search for TB treatment outcomes in Ethiopia, published between 2010 and 2025. Studies focused on specific factors, such as rural regions or treatment-resistance TB cases, were excluded (Limenh et al., 2024; Melese et al., 2016; Ereso et al., 2024; Wondale et al., 2017; Berhan et al., 2023; Biruk et al., 2016; Tsegaye et al., 2024; Ayele and Nenko, 2015; Alemu et al., 2021; Gebremariam et al., 2016; Debash et al., 2023; Tesfahuneygn et al., 2015).

### 2.3 Data Cleaning and Filtering

Our Botswana retrospective cohort included patients of any age with documented treatment outcomes for TB hospitals in Gaborone, Botswana. We excluded individuals with missing TB treatment outcome or incorrectly reported age or CD4+ T-cell counts. In the Pre-Treat All dataset, we only included patients from Gaborone with available TB treatment outcomes. We combined the Pre- and Post-Treat All datasets to perform analysis of the two eras. See Figure 1 for more details of our filtering steps.

**Figure 1.**
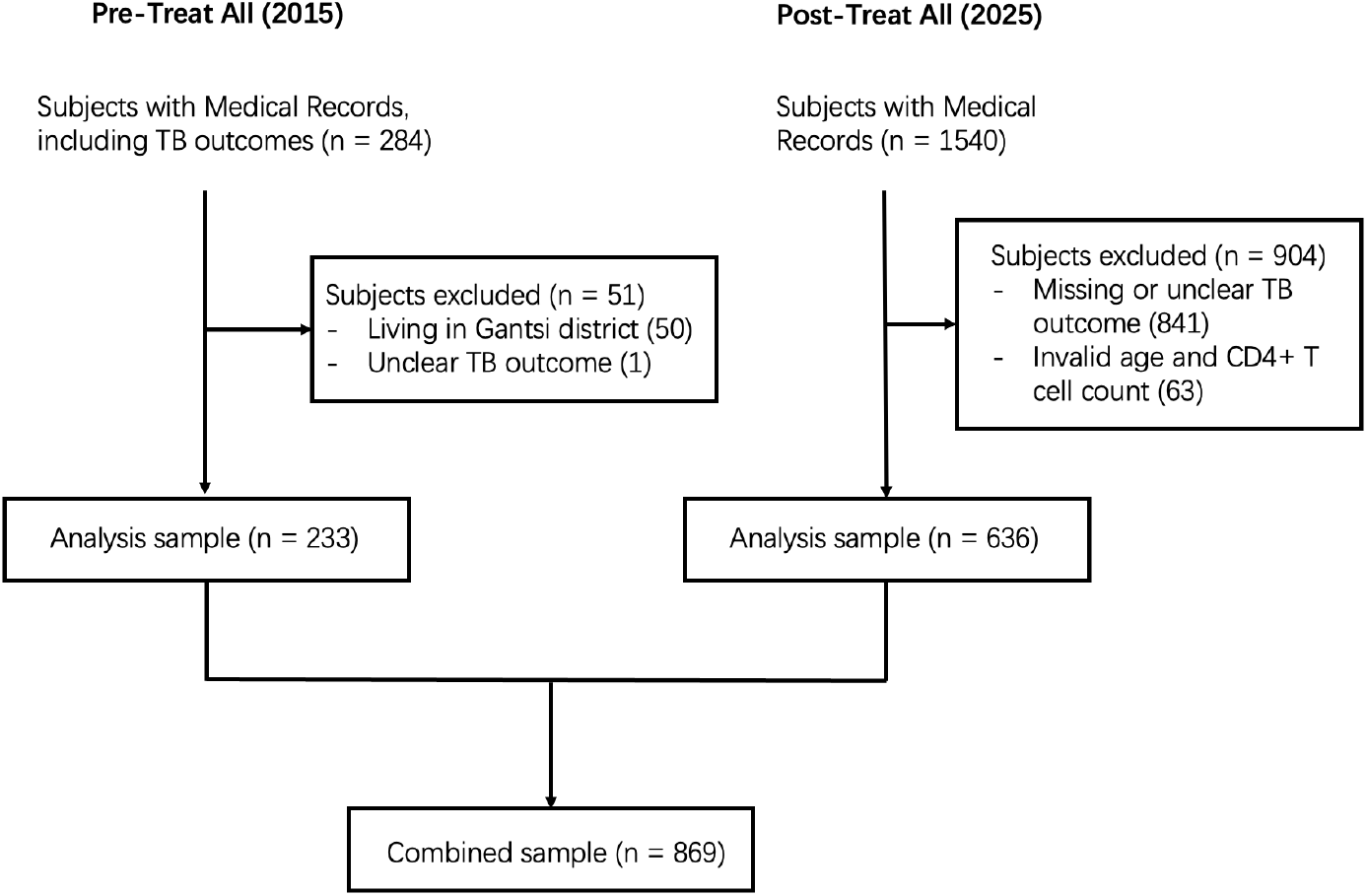
Flow chart showing data cleaning steps and sample sizes for analysis datasets.

### 2.4 Statistical Analysis

#### 2.4.1 Association betweeen HIV Status and TB Treatment Outcome

We first used logistic regression to quantify the relationship between the binary response variable, favorable/unfavorable TB treatment outcome, and the predictor of interest, HIV status:

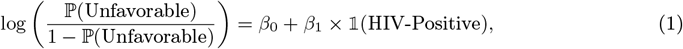

where 𝟙 is the indicator function such that 𝟙 (HIV-Positive) = 𝟙 for HIV positive individuals, and 𝟙 (HIV-Positive) = 0 otherwise. This model yields unadjusted inference on the relationship between HIV and unfavorable TB treatment outcomes in the Post-Treat All period. The parameter 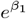 can be interpreted as the relative odds of an unfavorable TB treatment outcome, comparing the subpopulation of individuals with HIV to the subpopulation without HIV, unconditional on any other covariates.

We additionally fit an adjusted model, where covariates known to be related to the outcome were adjusted for; we included age (years), sex, distance between the patient’s home and clinic (km), substance use (smoking/consuming alcohol), prior history of TB, and occupational status:

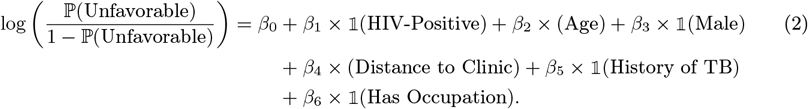

Model 2 gives adjusted odds ratios for the relationship between HIV infection and TB treatment outcome; 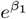 can now be interpreted as the relative odds of unfavorable TB treatment outcomes comparing the subpopulations with and without HIV who are similar in age, sex, distance to their clinic, TB history, and occupational status. The exclusion of ART history as a covariate is notable, since it is known to be part of the mechanism that changes the relationship between HIV status and TB treatment outcome. We deliberately excluded ART use as a covariate because it lies on the causal pathway between HIV infection and TB treatment outcome: ART use is a direct consequence of HIV status and mediates its biological effect on TB progression. Conditioning on ART history would therefore block part of the total effect of HIV that we wanted to infer. Instead, we interpret the coefficient for HIV status as capturing the total (direct and indirect) association between HIV infection and TB treatment outcomes in each era.

The patient’s level of immune function provides biological context for how HIV influences TB outcomes. To capture this, we consider CD4+ T-cell count, a marker of immune function that reflects HIV disease severity. We partitioned the HIV-positive covariate by CD4+ T-cell count groups (with cutoffs specified by WHO guidelines) to infer the relationship between immune function and TB treatment outcomes. This model with the stratified HIV covariate is

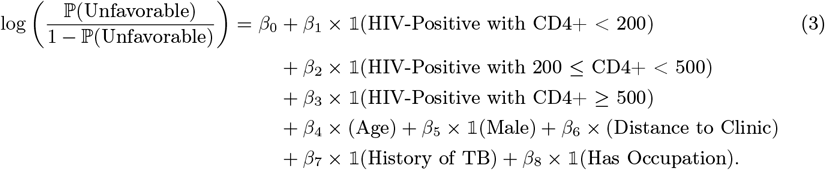

#### 2.4.2 Changes in the HIV Effect Between the Pre- and Post-Treat All Populations

To evaluate whether the relationship between HIV status and unfavorable TB treatment outcomes changed following Botswana’s universal ART policy, we used data from before the universal ART policy (Pre-Treat All) in addition to the recent data (Post-Treat All) and fit a simplified logistic regression model including an interaction between HIV status and study period:

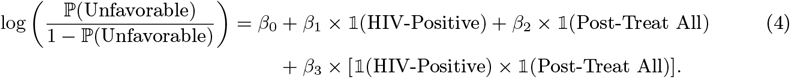

The coefficient of the interaction term, *β*_3_, captures the change in the effect of HIV status on TB outcomes between the two periods: a negative estimate of *β*_3_ would suggest that the disparity between HIV-positive and HIV-negative individuals narrowed after the introduction of universal ART. Because several covariates (e.g., occupation, CD4 count) were unavailable or inconsistently measured in the Pre-Treat All dataset, we fit this parsimonious model using only variables common to both datasets.

Given the limited sample size in the Pre–Treat All data, we anticipated imprecision in 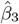 and extended the interaction-term analysis using a Bayesian mixed-effects model that incorporated the 2 Botswana studies and 12 Ethiopia studies described in Section 2.2. The model is defined as follows:

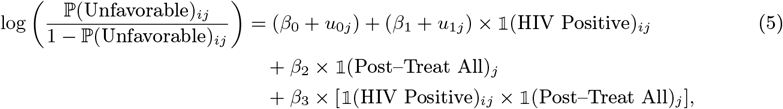

for individual *i* in study *j*. The fixed effects *β*_0_–*β*_3_ describe the population-average relationships, while *u*_0*j*_ and *u*_1*j*_ are zero-mean, normally distributed random effects (with variances 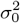 and 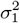,respectively) that allow the intercept and the HIV effect to vary across studies. Parameters were assigned weakly informative priors:

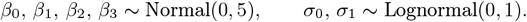

We report two related sets of results from this model. The pooled estimates reflect the population-level (fixed) effects, representing the average association between HIV status, treatment era, and outcome across all studies. The Botswana random-effects estimates incorporate the estimated study-specific deviations *u*_0*j*_ and *u*_1*j*_ for the two Botswana studies, combining evidence from all 14 datasets with information particular to Botswana through partial pooling. These estimates represent the posterior mean effects expected for the Botswana studies, conditional on both their observed data and the cross-study variation encoded in the hierarchical model.

#### 2.4.3 Statistical Software

We used R, version 4.5.1 (R Project for Statistical Computing; available at: http://www.r-project.org) for all statistical analyses performed in this study. Package `brms` (Bürkner, 2021, v2.22.0) was used to fit the Bayesian mixed effects model. In accordance with recent statistical guidelines, we did not specify a threshold for statistical significance.

#### 2.4.4 Ethical Considerations

The Pre-Treat All study was approved by the Institutional Review Boards (IRBs) at the US Centers for Disease Control and Prevention, University of California Irvine (UCI) and University of Pennsylvania, and the Botswana Ministry of Health Human Research Development Committee (HRDC). The Post-Treat-All study was approved by the IRBs at UCI and HRDC. All study participants provided written informed consent.

## 3 Results

In the Post-Treat All dataset, we excluded 841 individuals due to missing TB treatment outcome and 63 due to incorrectly reported age or CD4+ T-cell counts; this left us with a final sample size of 636. Descriptive statistics are given in Table 1. Among the Pre-Treat All comparison dataset, we excluded 50 individuals from cities outside of Botswana and one individual for unclear treatment outcome documentation, leading to a final Pre-Treat All sample size of 233.

**Table 1:**
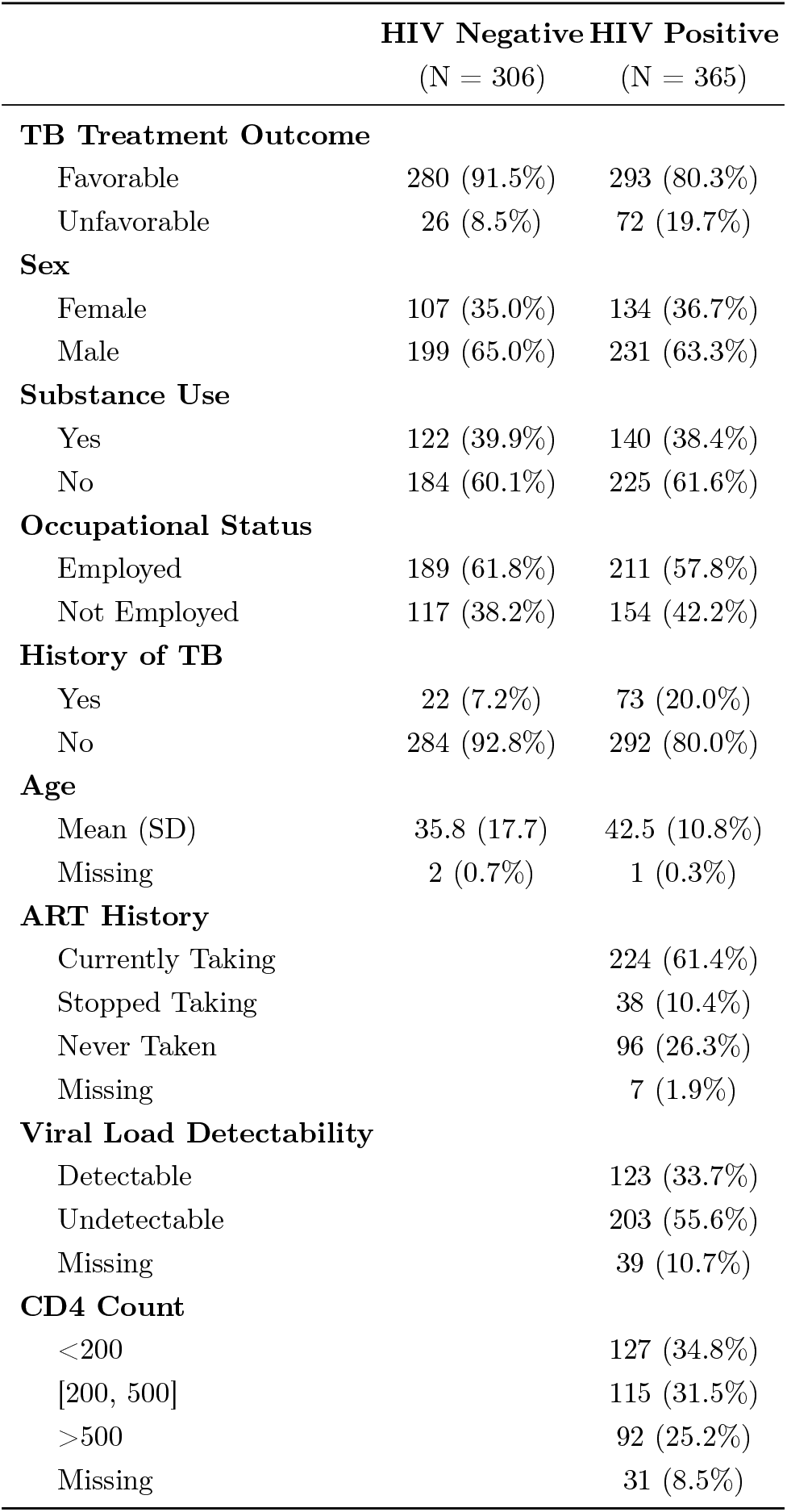
Descriptive statistics for all participants. ART History, Viral Load Detectability, and CD4 Counts are only applicable for HIV positive participants.

|  | **HIV Negative**<br>(N = 306) | **HIV Positive**<br>(N = 365) |
| --- | --- | --- |
| **TB Treatment Outcome** |  |  |
| Favorable | 280 (91.5%) | 293 (80.3%) |
| Unfavorable | 26 (8.5%) | 72 (19.7%) |
| **Sex** |  |  |
| Female | 107 (35.0%) | 134 (36.7%) |
| Male | 199 (65.0%) | 231 (63.3%) |
| **Substance Use** |  |  |
| Yes | 122 (39.9%) | 140 (38.4%) |
| No | 184 (60.1%) | 225 (61.6%) |
| **Occupational Status** |  |  |
| Employed | 189 (61.8%) | 211 (57.8%) |
| Not Employed | 117 (38.2%) | 154 (42.2%) |
| **History of TB** |  |  |
| Yes | 22 (7.2%) | 73 (20.0%) |
| No | 284 (92.8%) | 292 (80.0%) |
| **Age** |  |  |
| Mean (SD) | 35.8 (17.7) | 42.5 (10.8%) |
| Missing | 2 (0.7%) | 1 (0.3%) |
| **ART History** |  |  |
| Currently Taking |  | 224 (61.4%) |
| Stopped Taking |  | 38 (10.4%) |
| Never Taken |  | 96 (26.3%) |
| Missing |  | 7 (1.9%) |
| **Viral Load Detectability** |  |  |
| Detectable |  | 123 (33.7%) |
| Undetectable |  | 203 (55.6%) |
| Missing |  | 39 (10.7%) |
| **CD4 Count** |  |  |
| <200 |  | 127 (34.8%) |
| [200, 500] |  | 115 (31.5%) |
| >500 |  | 92 (25.2%) |
| Missing |  | 31 (8.5%) |

### 3.1 Finalized Combined Dataset

The final combined dataset consisted of 233 observations from the Pre-Treat All era dataset and 636 observations from the Post-Treat All dataset (Table 1). In the Pre-Treat All dataset, 87 HIV-negative and 135 HIV-positive individuals were present, and 307 HIV-negative and 368 HIV-positive individuals were present in the Post-Treat All dataset. Between datasets, we observed minor differences in the distributions of variables such as age, and CD4 counts, but no major changes were visible. HIV-negative individuals in the Pre-Treat All dataset had around 5 percent of higher outcomes compared to Post-Treat All.

### 3.2 Association between HIV Status and TB Treatment Outcome

In the bivariate logistic regression model (1), we found an odds ratio of 3.01 [95% CI: (1.96, 4.73)] for unfavorable treatment outcomes among HIV-positive participants compared to HIV-negative participants.

The effect of HIV was still high after adjustment for other covariates in the multivariable logistic regression (Table 2). The relative odds of an unfavorable TB treatment outcome, comparing HIV positive and HIV negative groups with similar values in the adjustment covariates, was estimated to be 2.51 [95% CI: (1.48, 4.38)]. There was additionally strong evidence for a relationship between occupational status and TB treatment outcome, with an odds ratio of 0.60 [95% CI: (0.36, 0.98)], or about 40% lower odds of an unfavorable outcome in the employed subpopulation.

**Table 2:**
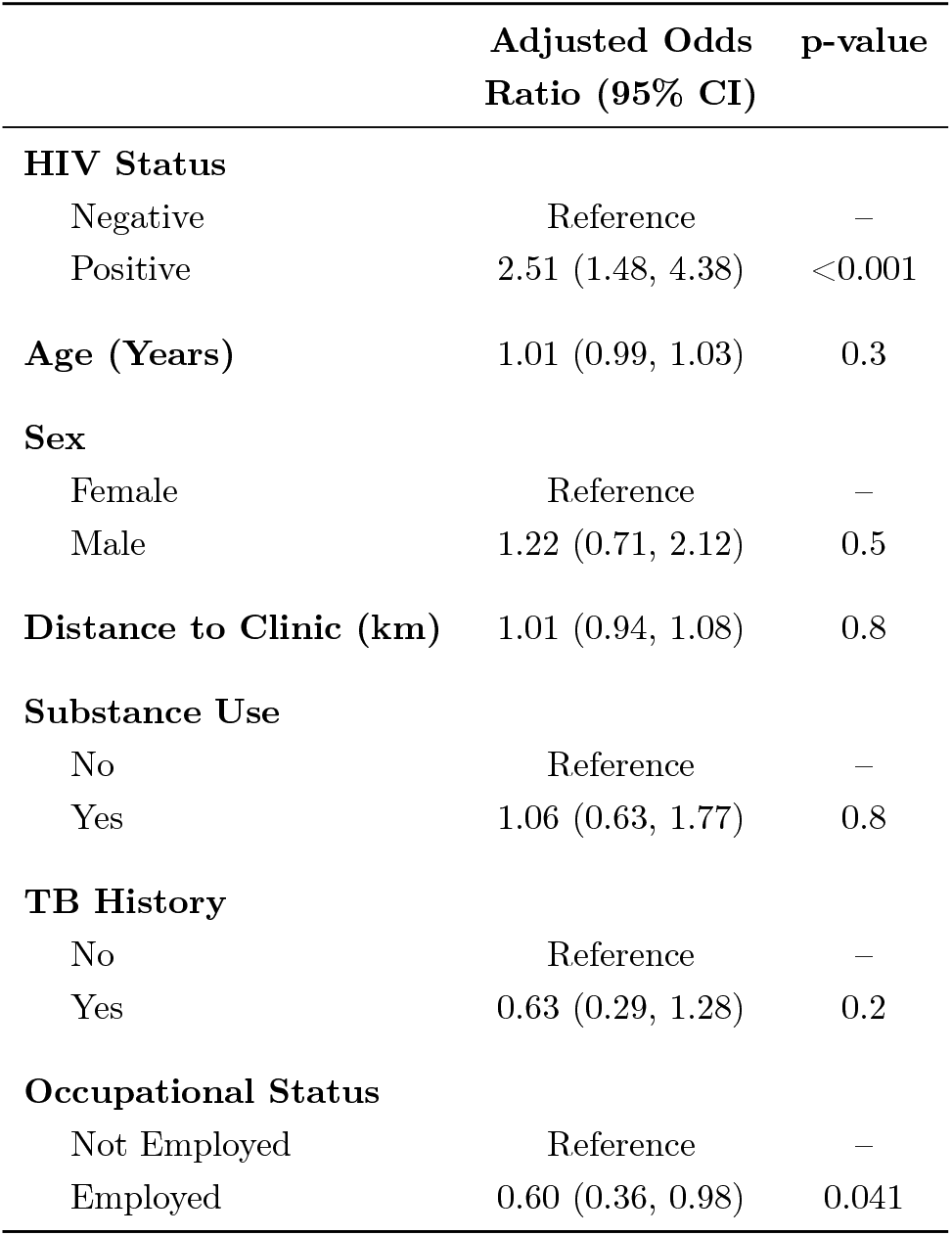
Odds ratios and associated 95% confidence intervals for Model 2. Odds ratios are reported on the unfavorable-outcome scale, such that values above one indicate higher odds of an unfavorable TB treatment outcome.

|  | Adjusted Odds<br>Ratio (95% CI) | p-value |
| --- | --- | --- |
| **HIV Status** |  |  |
| Negative | Reference | – |
| Positive | 2.51 (1.48, 4.38) | <0.001 |
| **Age (Years)** | 1.01 (0.99, 1.03) | 0.3 |
| **Sex** |  |  |
| Female | Reference | – |
| Male | 1.22 (0.71, 2.12) | 0.5 |
| **Distance to Clinic (km)** | 1.01 (0.94, 1.08) | 0.8 |
| **Substance Use** |  |  |
| No | Reference | – |
| Yes | 1.06 (0.63, 1.77) | 0.8 |
| **TB History** |  |  |
| No | Reference | – |
| Yes | 0.63 (0.29, 1.28) | 0.2 |
| **Occupational Status** |  |  |
| Not Employed | Reference | – |
| Employed | 0.60 (0.36, 0.98) | 0.041 |

Table 3 shows the results of Model 3 which partitions the HIV-positive condition by CD4+ T-cell counts. Compared to HIV-negative individuals, HIV-positive patients with CD4+ T-cell counts < 200 had over three times higher odds of an unfavorable outcome [OR: 3.12; 95% CI: (1.65, 5.97)]. There was some evidence that HIV-positive patients with CD4+ T-cell counts between 200 and 500 have higher odds of an unfavorable TB treatment outcome [OR: 2.04; 95% CI: (0.98, 4.19)] than HIV-negative patients, but no evidence that HIV-positive patients with CD4+ T-cell counts above 500 have higher odds of an unfavorable outcome [OR: 1.33; 95% CI: (0.55, 3.02)] as compared to HIV-negative patients.

**Table 3:** Odds ratios and associated 95% confidence intervals for Model 3. Odds ratios are reported on the unfavorable-outcome scale, such that values above one indicate higher odds of an unfavorable TB treatment outcome.

|  | Adjusted Odds<br>Ratio (95% CI) | p-value |
| --- | --- | --- |
| **HIV Status** |  |  |
| Negative | Reference | – |
| Positive with CD4+ > 500 | 1.33 (0.55, 3.02) | 0.5 |
| Positive with CD4+ in [200, 500] | 2.04 (0.98, 4.19) | 0.053 |
| Positive with CD4+ <200 | 3.12 (1.65, 5.97) | <0.001 |
| **Age (Years)** | 1.01 (0.99, 1.03) | 0.2 |
| **Sex** |  |  |
| Female | Reference | – |
| Male | 1.27 (0.72, 2.29) | 0.4 |
| **Distance to Clinic (km)** | 0.99 (0.91, 1.07) | 0.9 |
| **Substance Use** |  |  |
| No | Reference | – |
| Yes | 1.09 (0.63, 1.88) | 0.7 |
| **TB History** |  |  |
| No | Reference | – |
| Yes | 0.66 (0.29, 1.38) | 0.3 |
| **Occupational Status** |  |  |
| Not Employed | Reference | – |
| Employed | 0.56 (0.33, 0.94) | 0.029 |

### 3.3 HIV Effect in the Pre- and Post-Treat All Populations

Results from Models 4 and 5 are reported in Table 4, including point estimates with 95% confidence intervals for the frequentist analysis and with 95% Bayesian credible intervals (BCIs) for the Bayesian analysis. Inference from the frequentist interaction model, Model 4, was inconclusive. Using only the Botswana data, there was no evidence that the effect of HIV on TB treatment outcomes differed between the Pre-Treat All and Post-Treat All study periods [OR: 0.41; 95% CI (0.11, 1.55)].

**Table 4:** Comparison of odds ratios from the frequentist and Bayesian analyses. The frequentist results are based on a logistic regression using the two Botswana studies only. The Bayesian mixed-effects meta-analysis incorporates data from all 14 studies, with random effects on the intercept and HIV slope by study. The “pooled” Bayesian estimates represent population-level fixed effects averaged across studies, while the “Botswana random-effects” estimates additionally include the study-specific deviations for the two Botswana sites (one Pre-and one Post-Treat All). As before, odds ratios are reported on the unfavorable-outcome scale, such that values above one indicate higher odds of an unfavorable TB treatment outcome.

|  | **Frequentist Inference**<br>**(95% CI)** | **Bayesian Inference**<br>**(95% BCI)** |  |
| --- | --- | --- | --- |
|  | **Botswana studies only** | **All studies**<br>**pooled** | **Botswana studies**<br>**random-effects** |
| *Effect of HIV status (Positive vs. Negative):* |  |  |  |
| Pre-Treat All Period | 6.36 (1.86, 21.77) | 1.85 (1.28, 2.66) | 2.39 (1.36, 4.79) |
| Post-Treat All Period | 2.63 (1.63, 4.24) | 3.56 (2.35, 5.34) | 2.95 (1.95, 4.47) |
| *Effect of Study Period (Post- vs. Pre-Treat All):* |  |  |  |
| Among HIV- | 2.59 (0.77, 8.77) | 0.33 (0.16, 0.68) |  |
| Among HIV+ | 1.07 (0.65, 1.77) | 0.64 (0.27, 1.52) |  |
| *Interaction Effect (Relative Effect of HIV+ in Post- vs Pre-Treat All):* |  |  |  |
| Ratio of ORs | 0.41 (0.11, 1.55) | 2.39 (1.36, 3.34) | 1.22 (0.54, 2.54) |

Using the population-level (both Ethiopia and Botswana) inference from the Bayesian mixed effects model (Model 5), we found evidence that the negative effect of HIV on TB treatment outcomes was actually higher in the Post-Treat All period than in Pre-Treat All. The relative odds of an unfavorable TB treatment outcome, comparing HIV-positive and HIV-negative subpopulations, was estimated to be 2.39 times higher [95% BCI: (1.36, 3.34)] in the Post-Treat All period than in the Pre-Treat All period. This suggests that, on average across studies, the gap in outcomes by HIV status widened rather than narrowed.

Incorporating the Botswana-specific random effects, we found insufficient evidence of change in the effect of HIV status on unfavorably outcomes between the study periods: the relative odds ratio was estimated to be 1.22 [95% BCI: (0.54, 2.54)]. This estimate is more consistent with the Botswana-only frequentist analysis, which also found no evidence that the relative effect associated with HIV changed after the Treat All policy.

Beyond the interaction, Table 4 also shows that TB treatment outcomes likely improved overall in the period following the ART Treat All policy as compared to earlier. Among HIV-negative individuals, the pooled Bayesian model estimated a 67% reduction in the odds of an unfavorable outcome in the Post– versus Pre–Treat All period [OR: 0.33; 95% BCI: (0.16, 0.68)]. The same trend was observed in the point estimate for HIV-positive individuals [OR: 0.64; 95% BCI: (0.27, 1.52)], though with a smaller effect size, and the wide credible interval indicates considerable uncertainty about the magnitude and direction of this effect. Then, while the relative disadvantage associated with HIV may have persisted or increased modestly at the population level, absolute outcomes may have improved for both groups in the Post–Treat All era. This suggests that the apparent increase in the relative effect of HIV does not reflect worsening outcomes among HIV-positive patients. Rather, both HIV-positive and HIV-negative individuals may have experienced improvement in treatment outcomes in the period after implementation of the Treat All policy, but the evidence for improvement among HIV-positive individuals is weaker, with the 95% Bayesian credible interval spanning a wide range of plausible effects. The larger relative odds ratio for the effect of HIV on TB treatment outcomes comparing Post-vs. Pre-Treat All therefore reflects slower or less certain gains among HIV-positive patients, rather than a deterioration in their outcomes.

## 4 Discussion

Our results show that HIV infection remains a risk factor for unfavorable TB outcomes in Botswana during an era of universal access to ART. In addition, the proportion of TB outcomes was not found to have changed from the Pre-Treat All data set. Although Botswana achieved an impressive ART coverage rate after implementing the universal ART policy, HIV remains a significant risk factor for unfavorable TB treatment outcomes (Abdool Karim, 2016). A potential mechanism could be that prior to the Treat All policy, critically compromised HIV patients were already given access to ART. The Treat All policy, then, only extends ART to PLHIV with already normal or lightly compromised CD4+ T-cell counts and are already in a better position to combat Mtb infections, which leads to a marginal or insignificant effect on TB outcomes.

This study also shows that unemployment, which we interpret as a proxy for income, is also associated with unfavorable TB outcomes. This highlights the need to address disparities in the social determinants of health, alongside biomedical interventions.

Despite Botswana’s UNAIDS 95-95-95 achievement, the proportion of PLHIV in our data who have either never taken ART stopped taking ART is well above 5 percent. Thus, our study shows that among people with TB, a disproportionately high proportion of PLHIV are not on ART. Our overall results highlight that universal ART policies may not be quite enough to make PLHIV less vulnerable to unfavorable TB treatment outcomes. It is possible that the Treat All policy may fail to reach the most vulnerable people. Additional efforts should be made to effectively decrease the economic and logistic hurdles for PLHIV to obtain ART, while additional measures should be implemented to decrease the spread of HIV.

### 4.1 Limitations

We were not able to incorporate several key clinical biomarkers into our analysis, which may alter the effect size of our test results; these markers include viral load, TB drug resistance, and Mtb strain genotypes, the latter of which was previously proven in the Pre-Treat All dataset study to be strongly associated with TB treatment outcomes (Shin et al., 2018). Differences between the demographics of our two cohorts may impact our analysis. Our exploratory data analysis also shows that it is possible that there exists non-response bias among the participants, especially as being HIV positive is stigmatized in the society of Botswana. This could be addressed in the future with multiple imputation of missing data.

In light of the COVID-19 pandemic, which overlapped with the time frame of our study, we must consider the emergence of the SARS-CoV-2 virus starting in early 2019. SARS-CoV-2 can exacerbate Mtb-induced inflammatory conditions in the lung. Recent studies have already documented SARS-CoV-2 as a serious influence on TB incidence and mortality rates worldwide and predicts SARS-CoV-2 as a lingering threat for those at risk of TB (Adeyi and Oyeleke, 2017; Swartwood et al., 2025).

### 4.2 Concluding Remarks

Our analyses integrating health records, survey results, and clinical biomarkers demonstrates an association between HIV status and TB treatment outcomes among patients in Gaborone, Botswana in the Post-Treat All period. We also found that the relationship between HIV status and TB treatment outcomes remain like that of a decade ago, despite universal ART coverage on Botswana. Our study sheds light on the fact that PLHIV remain at higher risk, and that measures to spread awareness regarding TB prevention or to detect TB outbreaks are paramount in PLHIV. Future studies should be performed to incorporate further aspects of patient demographics, while considering new potential TB risk factors, such as SARS-CoV-2 and air pollution, as factors to be controlled. Future studies should also include rural communities, where factors such as pollution levels and common occupations may differ from urban regions.

## Data Availability

All data produced in the present study are available upon reasonable request to the authors

## 5 Acknowledgments

The authors would like to thank the study participants and data collection team in Gaborone, Botswana. This study was supported by funding from the US National Institute of Allergy and Infectious Diseases of the National Institutes of Health under awards no. R01AI147336, K01AI118559, R25AI170491, and R01AI097045.

